# Statistical design of Path to Prevention platform trial in early neuronal alpha-synuclein disease

**DOI:** 10.64898/2026.11.04.26367306

**Authors:** Barbara Wendelberger, Cora Allen-Savietta, Amy Crawford, Michael C. Brumm, Seung Ho Choi, Catherine M. Kopil, Ruth B. Schneider, Maggie M. Kuhl, David-Erick Lafontant, Dixie Ecklund, Kimberly Fabrizio, Tatiana Foroud, Frederic S. Goldstein, Cornelia Kamp, Karl Kieburtz, Danielle N. Larson, Nijee Luthra, Nicola Pavese, Andrew Siderowf, Craig E. Stanley, Caroline M. Tanner, Thomas F. Tropea, Tawny Willson, Sohini Chowdhury, Kenneth Marek, Tanya Simuni, Christopher S. Coffey, the Path to Prevention Investigators and Parkinson’s Precision Medicine Initiative

**Author notes:** Correspondence to: Barbara Wendelberger Berry Consultants, LLC, 3345 Bee Caves Rd, Suite 201, Austin, Texas 78746.

## Abstract

The Path to Prevention (P2P) platform trial is a first-in-class, phase 2A, randomized, double-blind, placebo-controlled learning study. Its design expedites the assessment of investigational therapies for the treatment of early-stage Neuronal α-Synuclein Disease (NSD), a biological disease continuum that encompasses and often precedes clinically manifest Parkinson’s Disease. P2P is embedded within the Parkinson’s Precision Medicine Initiative (PPMI), a robust observational longitudinal cohort study on Parkinson’s disease progression and a trial-facilitating enrollment framework.

A *regimen*, comprised of an active investigational therapy and a matched placebo control, evaluates a therapeutic candidate within the P2P perpetual platform framework. Multiple regimens can be enrolled, enabling efficient testing of multiple therapeutics. A two-stage randomization scheme determines regimen sample size with 125 participants assigned to a regimen’s active therapy and a variable number to placebo control. The treated follow up period is a minimum of 96 weeks and a maximum of 144 weeks, with the last randomized participant followed for 96 weeks.

As a pioneering interventional trial in early-stage NSD, P2P endpoint selection was critically informed by modeling and analysis of available PPMI data. In P2P, regimens will be evaluated for efficacy using two primary endpoints: 1) a biomarker outcome of Dopamine Transporter Single-Photon Emission Computed Tomography (DAT SPECT) imaging measured by the bilateral putamen Specific Binding Ratio (SBR) and 2) a clinical outcome measured by the Movement Disorder Society-Sponsored Revision of the Unified Parkinson’s Disease Rating Scale (MDS-UPDRS) Part III score. Each endpoint will be assessed separately using a longitudinal Bayesian mixed effects model, which compares the active treatment group to the shared placebo control group. The model uses regimen-specific controls at full weight and dynamically discounts controls from other regimens. Additionally, placebo control estimates are supplemented by non-randomized control data from the PPMI cohort.

Clinical trial simulations, informed by longitudinal data from 275 P2P-eligible PPMI study participants (Age 60-81, 35% Female, 65% Male), were conducted to evaluate the design. Each regimen is well-powered to detect a 40% slowing in progression for both the clinical and biomarker primary endpoints when comparing active treatment to randomized control data. Supplementing the control estimation with non-randomized, P2P-eligible registry participant data increases power in the explored scenarios up to 4% for each endpoint.

The embedded P2P platform trial is a powerful learning study that leverages statistical efficiencies from the platform trial design and established PPMI infrastructure and is poised to advance treatment development in early-stage NSD.

## Introduction

The Path to Prevention (P2P) platform trial is a first-in-class, phase 2A, randomized, double-blind, placebo-controlled learning study investigating multiple disease-modifying therapeutic candidates in participants with early stage, biologically defined Stage 2B Neuronal α-Synuclein Disease (NSD). α-Synuclein pathology is present in several neurodegenerative syndromes including Parkinson’s disease (PD) and dementia with Lewy bodies.^1–3^ The current treatment options that exist for early stage PD treat only symptoms and cannot slow or halt disease progression.^4–6^ Recent advances in a biological definition of NSD propose an integrated staging system (NSD-ISS) that captures the presence of pathological n-α-syn species and dopaminergic neuronal dysfunction, as well as the presence of clinical symptoms.^3^ With measurable biomarker pathology but minimal clinical symptoms, the Stage 2B NSD population is an ideal target for assessing biologically targeted therapeutics (BTTs) to slow or halt clinical progression.^7^ Narrowing the trial population to this biologically defined early stage of disease reduces heterogeneity in an effort to increase the likelihood of demonstrating treatment efficacy, a precision medicine strategy that has successfully been used in other neurodegenerative diseases.^8–11^ The companion manuscript to this paper (Schneider et al) further details the landscape of BTTs for PD and the rationale for using the NSD population for P2P.

P2P is a pioneering trial testing potential disease-modifying therapeutics in the early stages of the pathological NSD process, when participants exhibit limited manifestation of clinical symptoms. Endpoint selection for this novel trial population of biologically defined NSD Stage 2B participants required careful consideration. We surveyed the landscape of outcome measures used in PD trials, with particular focus on disease-modifying trials.^12,13^ The Movement Disorder Society-Sponsored Revision of the Unified Parkinson’s Disease Rating Scale (MDS-UPDRS) is a validated clinical outcome measure and is often considered a standard outcome assessment for PD.^14,15^ It has four Parts, which assess distinct domains of PD: non-motor experiences of daily living (Part I), motor experiences of daily living (Part II), motor examination (Part III), and motor complications (Part IV).^14^ Individual MDS-UPDRS Part scores, in particular Part III, as well as composite scores derived from individual MDS-UPDRS Part scores have been used as primary efficacy endpoints in PD clinical trials.^12,13,16–19^ MDS-UPDRS Part scores are clinimetrically distinct with differential longitudinal progression in early stages of disease.^14,20,21^ While these endpoint considerations in early PD may have some overlap with those for the Stage 2B NSD population, it was critically important to explore natural history study data to assess endpoints in the context of Stage 2B NSD, specifically. Outcome assessments of biological changes also play a role in PD trials, with Dopamine Transporter (DAT) imaging aiming to capture early dopaminergic neuronal dysfunction.^22^ As an example, the CALM-PD sub study used longitudinal DAT imaging as an outcome measure to quantify neuronal degeneration.^23^ In addition to capturing important biological information, DAT imaging captures pathological changes that may precede clinical changes.^24^ Selecting appropriate endpoints in early-stage NSD is critical to well-powered trial design.^25^

The P2P platform trial is embedded within the PPMI Clinical natural history study that is part of the longstanding Parkinson’s Precision Medicine Initiative (PPMI), which provides a robust and efficient infrastructure for identifying and enrolling trial-eligible participants (**Figure 1**).^26^ The PPMI natural history study contains longitudinal, observational data on the progression of numerous outcome measures across the stages of NSD. Within PPMI Clinical, participants receive in-clinic assessments and provide comprehensive longitudinal data on clinical, imaging, and biomarker endpoints. Participants are recruited into three phenotypically defined cohorts: Parkinson’s disease, Prodromal and Healthy controls.^27^ All participants undergo biological testing to assess the presence of neuronal α-synuclein and dopaminergic neuronal dysfunction, as well as extensive clinical characterization to define their NSD status (+/-). NSD+ participants are staged using the NSD-ISS. The Clinical cohort data provides valuable insight into the natural progression and associated variability of biological and clinical markers of NSD, including how these markers differ for NSD+ versus NSD-participants. These data were extensively modeled to inform P2P endpoint selection. In addition to facilitating trial recruitment, embedding P2P within the infrastructure that supports the PPMI Clinical cohort enables a key innovative design feature: the incorporation of non-randomized control data from P2P-eligible participants in the ongoing PPMI study to help characterize the natural progression of early-stage NSD. Randomized and non-randomized control data are collected under the same PPMI study umbrella and collected using the same standardized methods for data collection.

**Figure 1.**
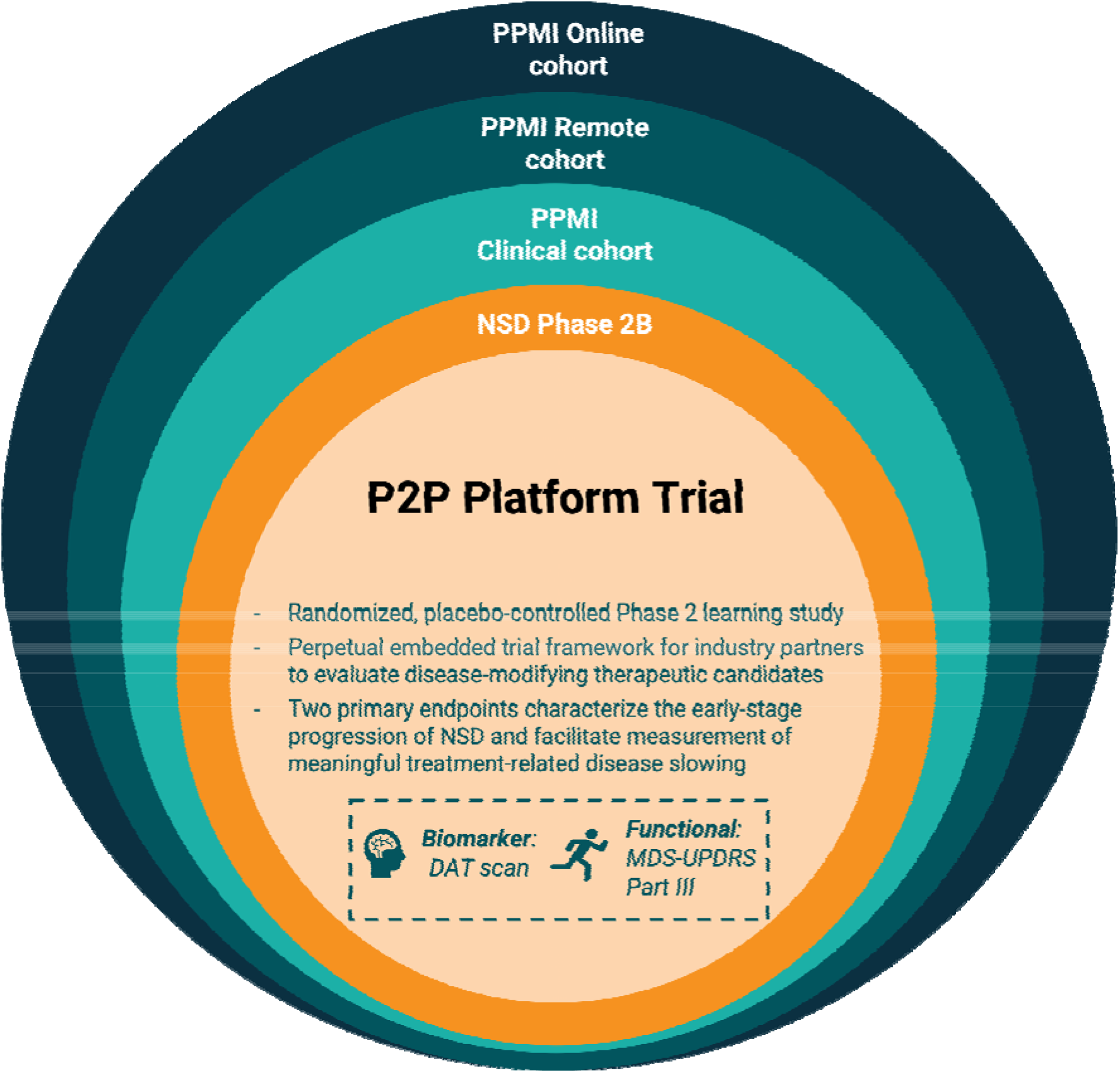
P2P platform trial embedded within PPMI. This figure displays the embedding of the P2P platform trial within the PPMI natural history study. Participants in the PPMI Online cohort provide patient-reported outcomes online and additional data from remote technologies, PPMI Remote serves as a tool for remote screening of population for hyposmia and other risk factors. These may include data from olfactory or genetic screening. Participants can move from the PPMI Remote cohort into the PPMI Clinical cohort, which includes in-person longitudinal data collection for clinical and imaging endpoints, and biospecimen sample collection. In the PPMI Clinical cohort, participants are classified using the NSD Staging system, and participants with NSD Stage 2B who are at least 60 years of age, do not have a clinical diagnosis of Parkinson’s disease, dementia with Lewy bodies or another neurodegenerative disease, and are not on dopaminergic therapy or a cholinesterase inhibitor are eligible for enrolment in P2P. The P2P platform trial is randomized, placebo controlled, and perpetual. Leveraging PPMI infrastructure, P2P supports therapeutic clinical development with both a biomarker and functional primary endpoint.

P2P provides a trial framework for industry partners to test therapeutic candidates within a *regimen*, which is comprised of an active investigational therapy and a matched placebo control. Multiple regimens can enroll in the platform at once, allowing the evaluation of multiple therapeutic candidates. The P2P primary analyses use longitudinal Bayesian mixed effects models to evaluate the disease progression rate for participants on active treatment versus shared placebo control as measured by the longitudinal primary endpoints, MDS-UPDRS Part III and DAT bilateral putamen specific binding ratio (SBR). We explored and refined statistical models using longitudinal data from participants in the PPMI Clinical cohort, selected to meet P2P inclusion criteria. Since P2P participants will be enrolled through PPMI, thorough analyses of these cohort data provided high-quality, evidence-based assumptions for platform trial simulation. With no prior precedent, novel endpoint exploration of the biologically defined trial population was a necessary part of the trial design process. Numerous trial simulations were conducted to evaluate the P2P platform trial design features and ensure a robust, well-powered, and efficient clinical trial design.

## Materials and methods

### Endpoint selection

We modeled PPMI natural history data to understand the rate, shape, and variability of disease progression in the NSD Stage 2B population, which has not previously been therapeutically targeted. Outcome assessments that capture early biological and clinical change are essential as endpoints in trials for early stage disease.^25^ Our team of clinicians and statisticians considered both the statistical properties and clinical value of various Dopamine Transporter Single-Photon Emission Computed Tomography (DAT SPECT) imaging outcomes, as well as MDS-UPDRS Total, Part I, Part III, and Part II+III scores, when selecting primary endpoints for the study. Statistical properties evaluated to assess outcome measures included sensitivity and specificity to phenoconversion, defined as investigator diagnosis of clinically defined neurodegenerative syndrome (ie Parkinson’s disease, Dementia with Lewy bodies and related conditions) as well as signal to noise ratio. In the PPMI data analyzed, the MDS-UPDRS Part III score had higher sensitivity to phenoconversion and higher signal to noise ratio than MDS-UPDRS Total score, while MDS-UPDRS Total score had higher specificity. Sensitivity and signal to noise ratio for MDS-UPDRS Part I and MDS-UPDRS Part II+III was lower than for MDS-UPDRS Part III. Clinician input on outcome assessment’s ability to capture relevant symptoms was also considered. Learnings from this statistical modeling, as well as clinician input, were used to support the P2P primary endpoint selection and primary analysis model specification. MDS-UPDRS Part III focuses on clinician-rated motor symptoms. Based on PPMI natural history study data analysis and the goal of a well-powered study, we chose to analyze MDS-UPDRS as a continuous endpoint. As a learning platform, P2P includes a wide range of exploratory endpoints. If any of these exploratory endpoints better capture change in the early NSD population, there is equipoise for updating the study to a more appropriate endpoint.

### Trial design

A key objective of the P2P platform trial is the ongoing ability to efficiently assess multiple therapeutic candidates for early-stage NSD. The P2P Master Protocol outlines a platform trial design that can accommodate several treatments at a time, each enrolled in a separate regimen (**Figure 2**).^28,29^ Each regimen contains a single active treatment and matched placebo control. Trial participants randomize in a two-step randomization process: 1) first with equal probability to a regimen for which they are eligible in an unblinded fashion, then 2) within that regimen, to receive active treatment or control with a K:1 ratio in a blinded fashion.^30^ Here, K denotes the number of actively enrolling regimens in the platform at the time of a participant’s randomization. When there is more than one active regimen, this allocation ratio increases the overall likelihood that an individual participant receives an active treatment while maintaining 1:1 allocation of each regimen’s active treatment participants to the platform’s randomized shared placebo controls. This two-step randomization process both enables the platform study design and requires careful statistical handling of control populations in the primary analysis. The pre-specified randomization scheme assigns 125 participants to active therapy and a variable number to placebo control. For participants randomized in P2P, follow up continues for a minimum of 96 weeks and a maximum of 144 weeks, with the last randomized participant followed for 96 weeks. A regimen’s primary analysis triggers 96 weeks after the last regimen participant is randomized. More details on the study design can be found in the companion manuscript by Schneider et al.

**Figure 2.**
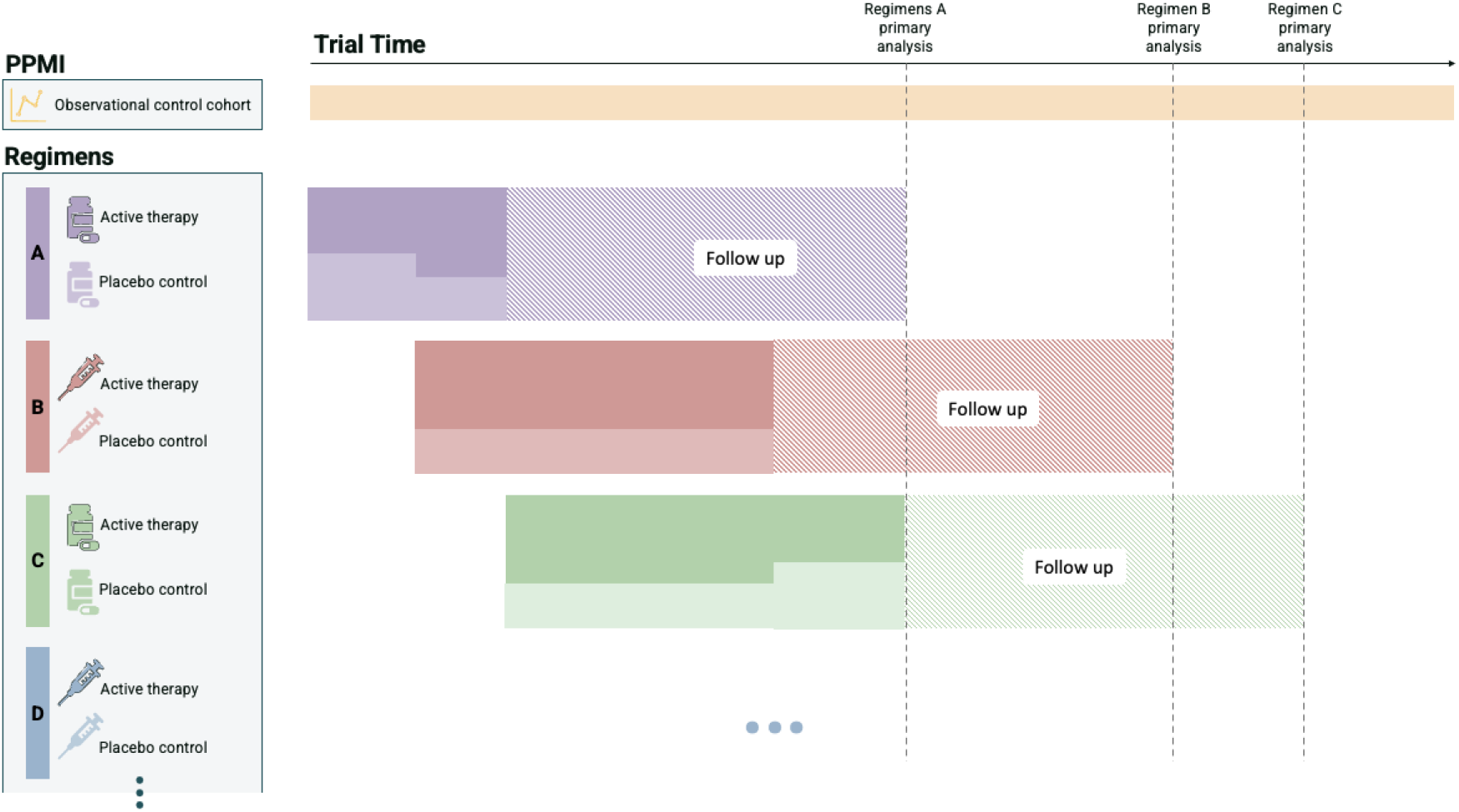
P2P platform trial design. This figure displays an example of the platform-level design of P2P. Regimens can enter the platform at different times and randomization to active therapy or placebo control, shown with shading, adjusts based on the number of actively enrolling regimens. The first regimen in the platform, Regimen A, is expected to enroll more quickly than later regimens, because of a bolus of participants identified through the PPMI study ready to enroll at the launch of P2P. Regimens enroll in the platform until 125 active therapy participants have been randomized. The primary analysis for a regimen is triggered 96 weeks after the last participant is randomized. Participants are followed until the time of their regimen’s primary analysis, or 144 weeks, whichever comes first. P2P-eligible participants in the PPMI observational cohort continue to accrue over time and are part of the collective control group used in the trial’s primary analysis. P2P is perpetual, with regimens entering and exiting the platform.

The P2P visit schedule encompasses the PPMI visit schedule. This longitudinal alignment enables natural incorporation of parallel observational data from participants in PPMI who are eligible but unwilling or unable to consent to enrollment in the P2P randomized trial. PPMI data collected on non-randomized PPMI participants supplements the randomized data collected on control participants in the trial to enable improved estimation of the natural progression of untreated participants during analysis. The statistical modeling framework discounts data from the non-randomized P2P-eligible participants in the PPMI Clinical cohort to account for systematic differences of these PPMI participants compared to the randomized placebo control group in P2P.

The P2P trial design employs two primary endpoints: 1) a biomarker outcome of DAT SPECT imaging as measured by the bilateral putamen Specific Binding Ratio (SBR) from baseline through follow-up, and 2) a clinical outcome measured by the MDS-UPDRS Part III score from baseline through end of follow-up or the initiation of dopaminergic treatment. Data for the DAT bilateral putamen SBR endpoint is collected at screening, 48 weeks, 96 weeks, and 144 weeks, and data for the MDS-UPDRS endpoint is collected at screening, baseline, 4 weeks, 12 weeks, 24 weeks, 48 weeks, 72 weeks, 96 weeks, 120 weeks, and 144 weeks. With the enrollment of Stage 2B NSD participants, the use of two primary endpoints aims to capture meaningful treatment-related disease slowing over time in the biomarker outcome, the clinical outcome, or both.

### Eligibility criteria

A participant is eligible for P2P if they are enrolled in PPMI, at least 60 years of age, NSD positive, and Stage 2B. To be included in NSD Stage 2B, participants must meet either a 1) cognitive, 2) motor, or 3) non-motor threshold. The 1) cognitive threshold is defined by Item 1.1 MDS-UPDRS ≥ 1 or Montreal Cognitive Assessment (MoCA) 25), ≥ 2) the motor threshold by subthreshold parkinsonism and MDS-UPDRS-II ≤ 2, and 3) the non-motor threshold by having REM sleep behavior disorder or hyposmia.^3^ Key exclusion criteria include diagnosis of PD or other neurodegenerative diseases, and participants cannot be taking PD medications at enrollment. Individual regimens may have additional eligibility requirements.

### Primary analysis population

The primary analysis population for a regimen includes an active treatment group and a collective control group (**Table 1**). When conducting a regimen’s primary analysis, the regimen of interest is referred to as the “analysis regimen.” The collective control group comprises intra-analysis regimen matched placebo controls (concurrent), extra-analysis regimen placebo controls (concurrent and nonconcurrent), and nonrandomized controls (NRC) from P2P-eligible participants in the PPMI Clinical cohort. Appropriate considerations are given to the composition of these control groups over time. A sensitivity analysis to the primary analysis population will be conducted restricting to concurrently randomized placebo control participants only.

**Table 1.** Analysis population definitions.

| Treatment group | Randomization group | Analysis regimen status | Name | Definition |
|---|---|---|---|---|
| Active therapy | Randomized | Intra-analysis regimen | Active treatment group | All participants randomized to active treatment in the analysis regimen |
| Collective Control | Randomized | Intra-analysis regimen | Intra-analysis regimen placebo controls | All participants randomized to control in the analysis regimen |
|  |  | Extra-analysis regimen | Extra-analysis regimen placebo controls | All participants randomized to control for a regimen other than the analysis regimen<br>Participants randomized during the period when the analysis regimen was being conducted are considered concurrent |
|  | Non-randomized | Extra-analysis regimen | Nonrandomized controls | Participants who were potentially eligible for P2P but refused or were unable to enrol and were followed up in the PPMI Clinical cohort |

### Handling control populations

The Bayesian modeling framework naturally enables borrowing strategies in the statistical design of the P2P platform trial.^29,31^ A dynamic borrowing strategy downweighs all extra-analysis regimen control data in a flexible way, borrowing from extra-regimen sources more heavily when the control data closely resembles the analysis regimen’s and with lower weight when the data are dissimilar. The collective control comprises distinct groups when considering enrollment status by regimen within three overarching categories (**Table 1**).

**1. Randomized Intra-Analysis Regimen Placebo Control Participants** Data is used with full weight in the primary analysis models. Placebo control participants are enrolled and randomized in the analysis regimen.
**2. Randomized Extra-Analysis Regimen Placebo Control Participants** Control participants from extra-analysis regimens are expected to have similar NSD progression to those in the analysis regimen, although each primary analysis model allows for deviations from this expectation. For example, if an extra-analysis regimen has a different mode of administration (e.g. injection vs oral) than the analysis regimen, it is possible that placebo effects may differ. We expect potential differences to be quite small in the biomarker endpoint, which is an objective measure expected to be robust to placebo effects. In the clinical MDS-UPDRS Part III endpoint, placebo effects have a greater potential to occur and to a greater magnitude. Given this, the selected prior for the DAT model promotes borrowing and downweighs only slightly. Conversely, the prior selected for the MDS-UPDRS Part III model is more flexible, allowing for strong downweighing of extra-analysis regimen controls, as appropriate, to account for the increased likelihood of differential placebo effects for the clinical endpoint.

a. **Concurrent** These participants are enrolled in a regimen or regimens other than the analysis regimen during the same time that the analysis regimen is active in the platform.
b. **Nonconcurrent** These participants are enrolled in a regimen or regimens other than the analysis regimen prior to the time that the analysis regimen is active in the platform.
3. **Nonrandomized Extra-Analysis Regimen Control Participants** These participants are P2P-eligible participants from the PPMI Clinical cohort who declined or were unable to randomize into P2P. While we expect non-randomized and randomized control participants in the NSD Stage 2B population to have similar NSD progression as measured by the primary endpoints, the primary analysis models allow for possible variability. These control participants are downweighed in the analysis through the same dynamic borrowing framework described in 2. Randomized Extra-Analysis Regimen.

In P2P, extra-analysis regimen control data shares substantial data collection infrastructure and has a much higher degree of similarity to the intra-analysis regimen data than external or historical control data.

### Primary analysis models

Treatment efficacy of the analysis regimen is assessed using a separate longitudinal Bayesian mixed-effects model for each primary endpoint (**Table 2 & Table 3)**. The Bayesian modeling framework was developed with clinical and statistical input and incorporates prior knowledge about endpoint progression through diffuse prior distributions. The prior distributions are centered at observed data values (means, variances, and correlations) from PPMI participants who are representative of the P2P trial population.

**Table 2.**
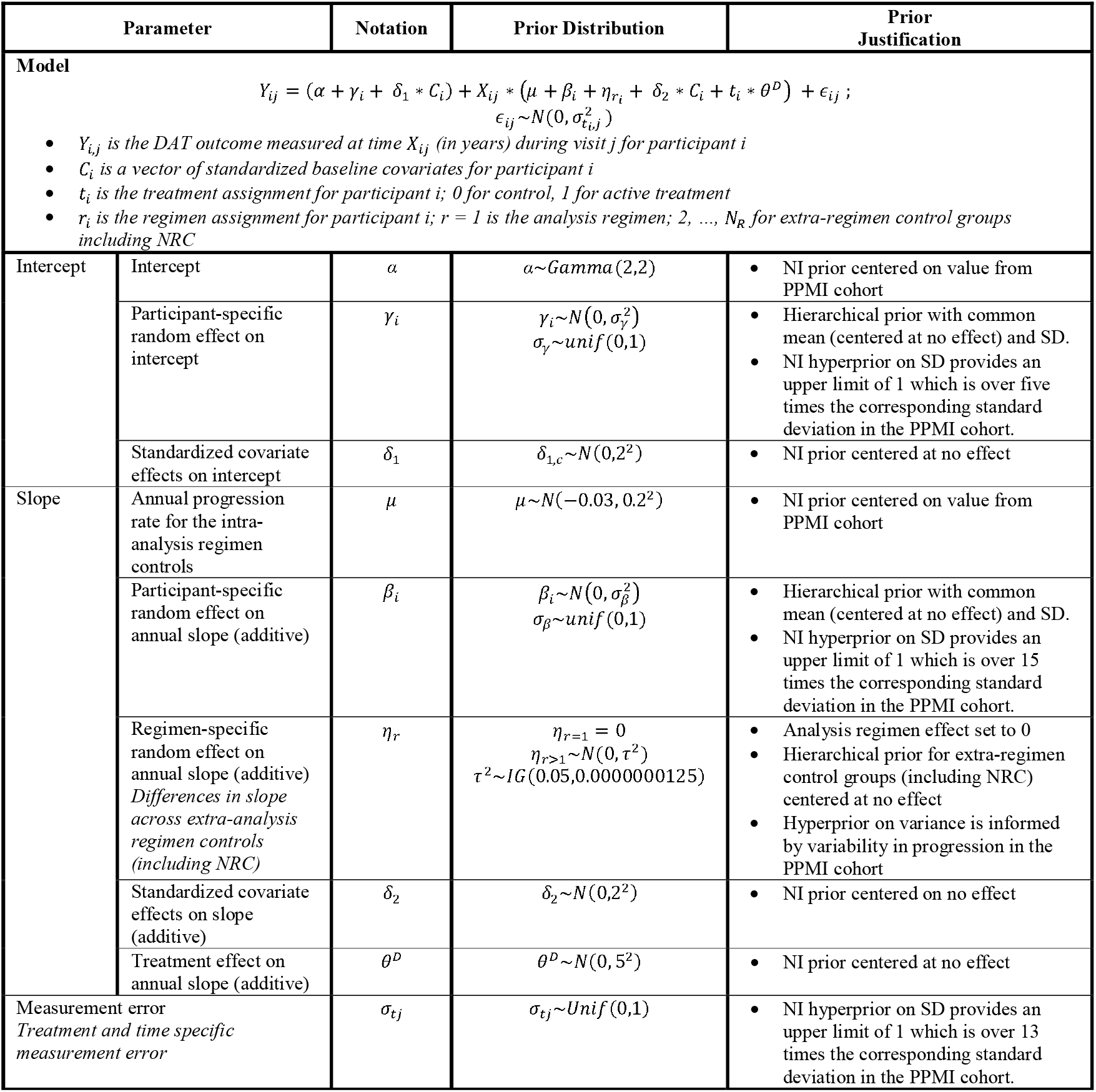
Primary analysis model for DAT bilateral putamen SBR. (DAT = DAT bilateral putamen SBR; SD = standard deviation; NRC = non-randomized controls; NI = non-informative)

| Parameter |  | Notation | Prior Distribution | Prior<br>Justification |
|---|---|---|---|---|
| **Model**<br>Y<sub>ij</sub> = (α + γ<sub>i</sub> + δ<sub>1</sub> \* C<sub>i</sub>) + X<sub>ij</sub> \* (μ + β<sub>i</sub> + η<sub>r<sub>i</sub></sub> + δ<sub>2</sub> \* C<sub>i</sub> + t<sub>i</sub> \* θ<sup>D</sup>) + ϵ<sub>ij</sub>;<br>ϵ<sub>ij</sub>~N(0, σ<sup>2</sup><sub>t<sub>i,j</sub></sub>)<br>• *Y<sub>i,j</sub> is the DAT outcome measured at time X<sub>ij</sub> (in years) during visit j for participant i*<br>• *C<sub>i</sub> is a vector of standardized baseline covariates for participant i*<br>• *t<sub>i</sub> is the treatment assignment for participant i; 0 for control, 1 for active treatment*<br>• *r<sub>i</sub> is the regimen assignment for participant i; r = 1 is the analysis regimen; 2, …, N<sub>R</sub> for extra-regimen control groups including NRC* |  |  |  |  |
| Intercept | Intercept | α | α~Gamma(2,2) | • NI prior centered on value from PPMI cohort |
|  | Participant-specific random effect on intercept | γ<sub>i</sub> | γ<sub>i</sub>~N(0, σ<sub>γ</sub><sup>2</sup>)<br>σ<sub>γ</sub>~unif(0,1) | • Hierarchical prior with common mean (centered at no effect) and SD.<br>• NI hyperprior on SD provides an upper limit of 1 which is over five times the corresponding standard deviation in the PPMI cohort. |
|  | Standardized covariate effects on intercept | δ<sub>1</sub> | δ<sub>1,c</sub>~N(0,2<sup>2</sup>) | • NI prior centered at no effect |
| Slope | Annual progression rate for the intra-analysis regimen controls | μ | μ~N(−0.03, 0.2<sup>2</sup>) | • NI prior centered on value from PPMI cohort |
|  | Participant-specific random effect on annual slope (additive) | β<sub>i</sub> | β<sub>i</sub>~N(0, σ<sub>β</sub><sup>2</sup>)<br>σ<sub>β</sub>~unif(0,1) | • Hierarchical prior with common mean (centered at no effect) and SD.<br>• NI hyperprior on SD provides an upper limit of 1 which is over 15 times the corresponding standard deviation in the PPMI cohort. |
|  | Regimen-specific random effect on annual slope (additive)<br>*Differences in slope across extra-analysis regimen controls (including NRC)* | η<sub>r</sub> | η<sub>r=1</sub> = 0<br>η<sub>r>1</sub>~N(0, τ<sup>2</sup>)<br>τ<sup>2</sup>~IG(0.05,0.0000000125) | • Analysis regimen effect set to 0<br>• Hierarchical prior for extra-regimen control groups (including NRC) centered at no effect<br>• Hyperprior on variance is informed by variability in progression in the PPMI cohort |
|  | Standardized covariate effects on slope (additive) | δ<sub>2</sub> | δ<sub>2</sub>~N(0,2<sup>2</sup>) | • NI prior centered on no effect |
|  | Treatment effect on annual slope (additive) | θ<sup>D</sup> | θ<sup>D</sup>~N(0, 5<sup>2</sup>) | • NI prior centered at no effect |
| Measurement error<br>*Treatment and time specific measurement error* |  | σ<sub>tj</sub> | σ<sub>tj</sub>~Unif(0,1) | • NI hyperprior on SD provides an upper limit of 1 which is over 13 times the corresponding standard deviation in the PPMI cohort. |

**Table 3.**
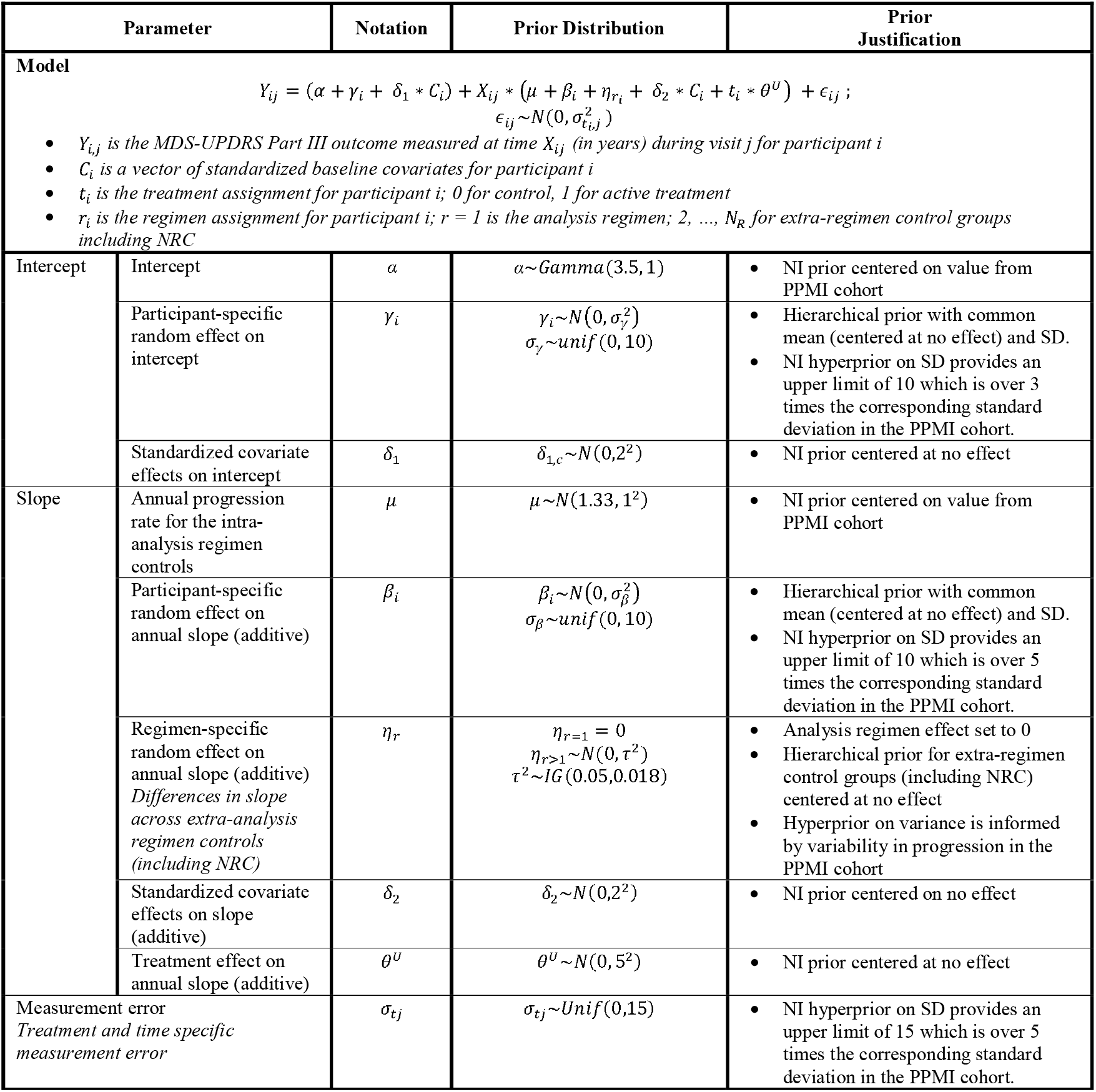
Primary analysis model for MDS-UPDRS Part III. (SD = standard deviation; NRC = non-randomized controls; NI = non-informative)

| Parameter |  | Notation | Prior Distribution | Prior<br>Justification |
|---|---|---|---|---|
| **Model**<br>Y<sub>ij</sub> = (α + γ<sub>i</sub> + δ<sub>1</sub> \* C<sub>i</sub>) + X<sub>ij</sub> \* (μ + β<sub>i</sub> + η<sub>r<sub>i</sub></sub> + δ<sub>2</sub> \* C<sub>i</sub> + t<sub>i</sub> \* θ<sup>U</sup>) + ϵ<sub>ij</sub>;<br>ϵ<sub>ij</sub>~N(0, σ<sup>2</sup><sub>t<sub>i,j</sub></sub>)<br>• *Y<sub>i,j</sub> is the MDS-UPDRS Part III outcome measured at time X<sub>ij</sub> (in years) during visit j for participant i*<br>• *C<sub>i</sub> is a vector of standardized baseline covariates for participant i*<br>• *t<sub>i</sub> is the treatment assignment for participant i; 0 for control, 1 for active treatment*<br>• *r<sub>i</sub> is the regimen assignment for participant i; r = 1 is the analysis regimen; 2, …, N<sub>R</sub> for extra-regimen control groups including NRC* |  |  |  |  |
| Intercept | Intercept | α | α~Gamma(3.5, 1) | • NI prior centered on value from PPMI cohort |
|  | Participant-specific random effect on intercept | γ<sub>i</sub> | γ<sub>i</sub>~N(0, σ<sub>γ</sub><sup>2</sup>)<br>σ<sub>γ</sub>~unif(0, 10) | • Hierarchical prior with common mean (centered at no effect) and SD.<br>• NI hyperprior on SD provides an upper limit of 10 which is over 3 times the corresponding standard deviation in the PPMI cohort. |
|  | Standardized covariate effects on intercept | δ<sub>1</sub> | δ<sub>1,c</sub>~N(0,2<sup>2</sup>) | • NI prior centered at no effect |
| Slope | Annual progression rate for the intra-analysis regimen controls | μ | μ~N(1.33, 1<sup>2</sup>) | • NI prior centered on value from PPMI cohort |
|  | Participant-specific random effect on annual slope (additive) | β<sub>i</sub> | β<sub>i</sub>~N(0, σ<sub>β</sub><sup>2</sup>)<br>σ<sub>β</sub>~unif(0, 10) | • Hierarchical prior with common mean (centered at no effect) and SD.<br>• NI hyperprior on SD provides an upper limit of 10 which is over 5 times the corresponding standard deviation in the PPMI cohort. |
|  | Regimen-specific random effect on annual slope (additive)<br>*Differences in slope across extra-analysis regimen controls (including NRC)* | η<sub>r</sub> | η<sub>r=1</sub> = 0<br>η<sub>r>1</sub>~N(0, τ<sup>2</sup>)<br>τ<sup>2</sup>~IG(0.05,0.018) | • Analysis regimen effect set to 0<br>• Hierarchical prior for extra-regimen control groups (including NRC) centered at no effect<br>• Hyperprior on variance is informed by variability in progression in the PPMI cohort |
|  | Standardized covariate effects on slope (additive) | δ<sub>2</sub> | δ<sub>2</sub>~N(0,2<sup>2</sup>) | • NI prior centered on no effect |
|  | Treatment effect on annual slope (additive) | θ<sup>U</sup> | θ<sup>U</sup>~N(0, 5<sup>2</sup>) | • NI prior centered at no effect |
| Measurement error<br>*Treatment and time specific measurement error* |  | σ<sub>tj</sub> | σ<sub>tj</sub>~Unif(0,15) | • NI hyperprior on SD provides an upper limit of 15 which is over 5 times the corresponding standard deviation in the PPMI cohort. |

Each primary analysis model is a longitudinal Bayesian mixed-effects model, which assumes a linear rate of progression over the period of participant follow-up. Each model incorporates participant-level random effects in baseline value of outcomes (intercept) & rate of progression (slope). The biomarker primary analysis model compares DAT bilateral putamen SBR in participants randomized to the analysis regimen’s treatment arm to participants in the collective control group. Similarly, the clinical primary analysis model compares MDS-UPDRS Part III progression rates in participants randomized to the analysis regimen’s treatment arm to participants in the collective control group. For the MDS-UPDRS Part III comparison, participants are censored at the initiation of any dopaminergic therapy though expected rate of such is below 5% based on PPMI data.

Treatment benefit for each endpoint is measured separately as the difference in annual progression rate, *θ^D^* and *θ^U^*, for DAT bilateral putamen SBR and MDS-UPDRS Part III, respectively. As shown in **Equation (1)**, a regimen demonstrates treatment benefit if the posterior probability that the treated participants progress more slowly than the control participants is greater than 0.90 for either endpoint.

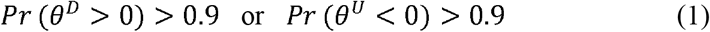

The treatment effect for DAT bilateral putamen SBR will also be reported as a progression rate ratio, PRR_DAT_, where the estimated additive treatment effect, *θ^^D^* is translated to a multiplicative effect on the estimated annual rate of progression for the control group in the analysis regimen, *μ^^D^*.

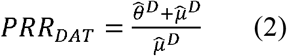

A treatment that slows progression will have a PRR less than one, indicating that participants on active treatment have slower disease progression than those in the collective control. For example, *PRR_DAT_* = 0.7 corresponds to a 30% estimated slowing over time in the rate of DAT decline for active treatment relative to control.

The treatment effect for MDS-UPDRS Part III is reported similarly.

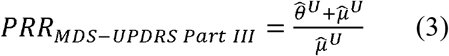

### PPMI Clinical cohort analysis

Accurate characterization of the natural progression of disease has important ramifications for modeling and trial design, and, subsequently, for trial operating characteristics such as power and Type I error. We analyzed data from the PPMI Clinical cohort to characterize important biomarker and clinical features of NSD Stage 2B progression (**See Supplemental Material**).^32^ Estimates from these models (**Supplemental Table S1**) were used to support simulations that inform clinical trial design and allow characterization of the trial’s operating characteristics.

### Clinical trial simulation

Clinical trial simulation is a powerful trial design tool used extensively in the development of the P2P Master Protocol. Accurate characterization of untreated disease progression based on data from the PPMI Clinical cohort provides robust estimates of endpoint trajectories and associated variability that are used to underpin trial simulation assumptions. Simulation assumptions for the primary endpoints as well as operational trial features such as accrual rate and dropout rate were evaluated by clinical and operational experts. The interplay of clinical, statistical, and operational expertise was critical to designing P2P, as the early-stage NSD population is only now being systematically characterized.^33^

Natural history study estimates serve as the basis for virtual participant simulation. A participant simulator creates realistic endpoint trajectories for DAT bilateral putamen SBR and MDS-UPDRS Part III outcomes for virtual P2P participants. Simulated control participant trajectories are centered around the estimates provided in **Supplemental Table S1**. Imposing a treatment effect in the simulation model then enables simulation of data for active treatment participants. As in the real trial, virtual participants are randomized to a regimen and then subsequently assigned to active treatment or matched placebo control. Primary endpoints for each virtual participant are simulated across time conditional on their regimen treatment assignment.

Covariance structures across time and endpoints were informed by analyses of the PPMI Clinical cohort data (**Supplemental Table S2**). Simulated participant data was generated by sampling multivariate standard normal outcomes for each participant for both longitudinal endpoints incorporating correlations within and between endpoints across time. Then, the standard normal deviates were transformed to the data scale by multiplying by the assumed standard deviation and then adding the assumed mean.

Simulated participant recruitment is assumed to be fast in the first year (300 participants per year), reflecting the P2P-ready participants already enrolled in the PPMI Clinical cohort at the outset of the P2P trial. In subsequent years, recruitment will rely on newly enrolled PPMI participants resulting in a slower rate (assumed to be 175 participants per year). Here, we assume regimen entry times of year 0, 0.5, 1, 1.8 years for regimens one through four, respectively.

Thousands of simulated trials were conducted with variations in sample size, untreated progression rate, data variability, regimen entry times, participant enrollment patterns, follow-up duration, randomization scheme, shared controls, and analysis strategies. Operating characteristics, averaged over many simulations, enabled the quantification and assessment of design performance and were used to iterate on design features before selecting a final design. These operating characteristics include trial power and Type I error rate. Clinical trial simulation was conducted in R (R Core Team 2025). **Supplemental Table S3** presents varying rates of control progression and their associated impact on trial operating characteristics.

## Results

### Clinical trial simulation

Clinical trial simulation enabled the simulation of thousands of virtual trials to assess design behavior and operating characteristics. A single virtual example trial highlights how the design and primary analysis function. **Figure 3** depicts the Regimen 2 primary analysis results from a single simulation of the P2P platform trial to illustrate expected data patterns and variability, how regimens enter the platform, their effect on randomization, and the accumulation of collective control information.

**Figure 3.**
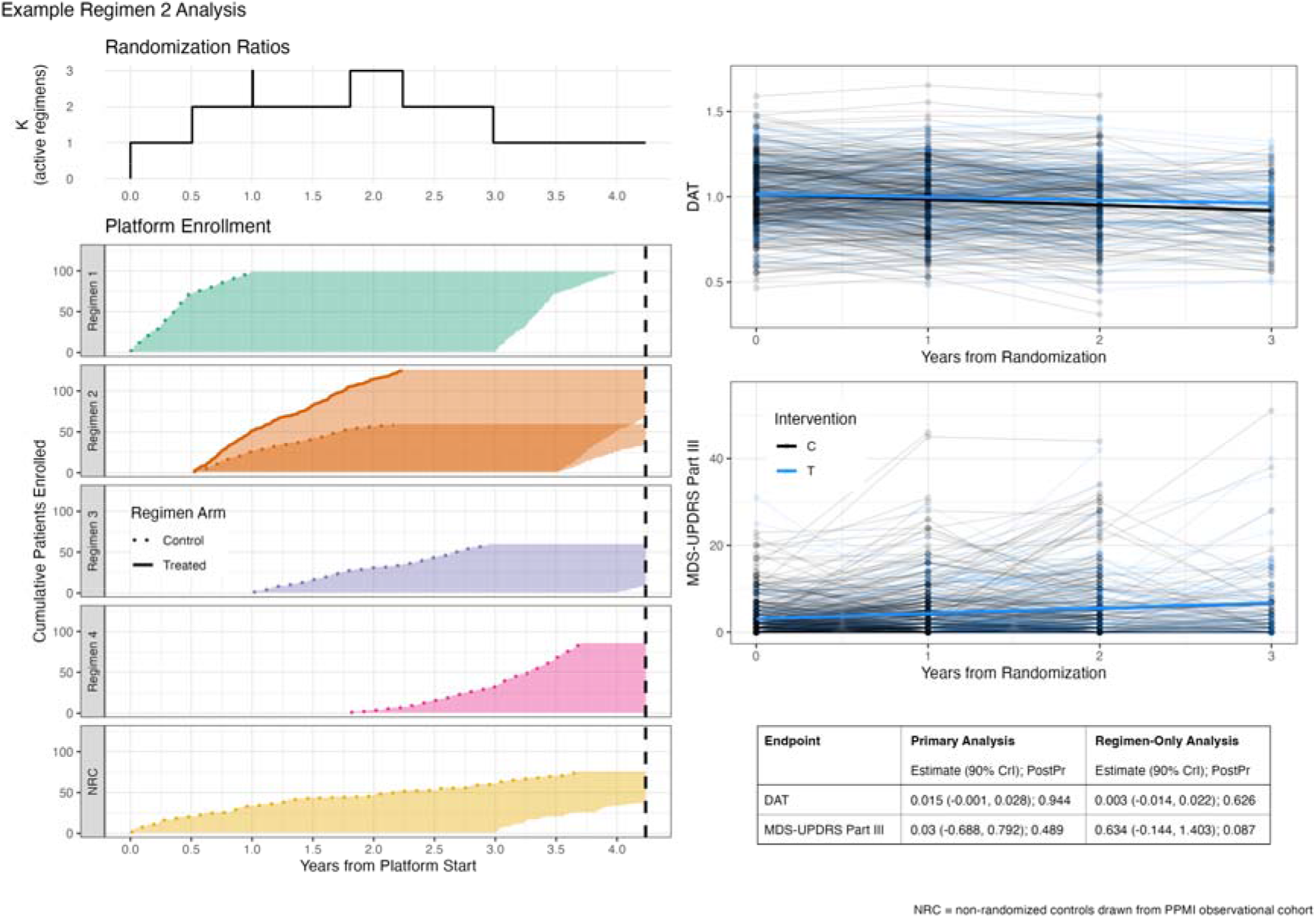
Simulation of P2P Platform Trial: Regimen 2 Analysis. This multi-panel figure demonstrates how the P2P design functions during the primary analysis of Regimen 2 using a single simulated data set. The top left panel titled Randomization Ratios shows how the K:1 within-regimen randomization varies as a function of the number of actively enrolling regimens in the platform (K). The bottom left panel titled Platform Enrollment shows the cumulative enrollment of participants across regimens, where each regimen is color coded and contained in a separate facet of the graph. Because this figure highlights the Regimen 2 analysis, active treatment participants are only shown in for Regimen 2, while the collective control group is comprised of available data from all regimens, as well as the non-randomized control (NRC). The upper right panel provides a participant-level spaghetti plot, as well as model-estimated mean trajectories for both the Regimen 2 active treatment group and the collective control group for the DAT bilateral putamen endpoint. The posterior probability that the DAT slope for the active treatment group shows less progression than the DAT bilateral putamen slope for the collective control is 0.94, which meets the pre-specified threshold for statistical significance. The lower right plane shows the MDS-UPDRS Part III spaghetti plot and model estimated mean trajectories. The posterior probability that the MDS-UPDRS Part III slope for the active treatment group shows less progression than the DAT bilateral putamen slope for the collective control is 0.63, which does not meet the pre-specified threshold for statistical significance. In this example trial, the primary analysis supports the conclusion of statistically significant slowing on the DAT bilateral putamen endpoint but not on the MDS-UPDRS Part III endpoint.

The top left panel highlights the K:1 active treatment to placebo control randomization, which varies based on the number of actively enrolling regimens in the platform. When regimen 1 enters the platform at time 0, within regimen randomization is 1:1. When regimen 2 enters the platform at time 0.5, the within regimen randomization changes to 2:1. Randomization increases to 3:1 when there are 3 actively enrolling regimens and decreases as regimens complete follow up and exit the platform. Cumulative participant enrollment is displayed in the bottom left panel with participant enrollment corresponding to the time each regimen is active in the platform. Collective control participants are shown across all regimens, as well as the nonrandomized cohort (NRC). Treated participants are shown only for regimen 2, as this example trial highlights the regimen 2 analysis.

The right column of the figure shows participant spaghetti plots and primary analyses for both DAT bilateral putamen and MDS-UPDRS Part III endpoints. Treatment effect estimates, as well as corresponding posterior probabilities, are shown for each endpoint in the table found at the bottom right. In this simulated example, the primary analysis for DAT successfully slows progression in the treated participants relative to the collective control with a posterior probability (0.944) greater than the pre-specified threshold of 0.9. The sensitivity analysis using regimen-only controls for DAT also suggests a treatment effect (0.003) but the posterior probability (0.626) does not reach the prespecified threshold for success. The MDS-UPDRS Part III endpoint does not demonstrate statistically significant slowing of progression in this simulated example.

### Handling control populations

The collective control design allows each regimen to draw on control participants across the platform trial, which increases the statistical efficiency and reduces the number of participants randomized to placebo control relative to a set of traditional two-arm placebo-controlled clinical trials. The primary analysis models employ a dynamic borrowing framework to handle potential heterogeneity in control progression rates across data different sources. **Figure 4** quantifies the impacts of heterogeneous control progression by setting the regimen 2 control participants to progress 15% slower or faster, on average, than all other control participants in the platform trial. We use 15% to benchmark behavior because it corresponds to approximately one standard deviation for the mean annual progression rate of each endpoint estimated from the PPMI natural history data. The figure presents operating characteristics demonstrating design behavior in the case of a discrepant control progression rate in regimen 2 and an expected control progression rate for regimen 3.

**Figure 4.**
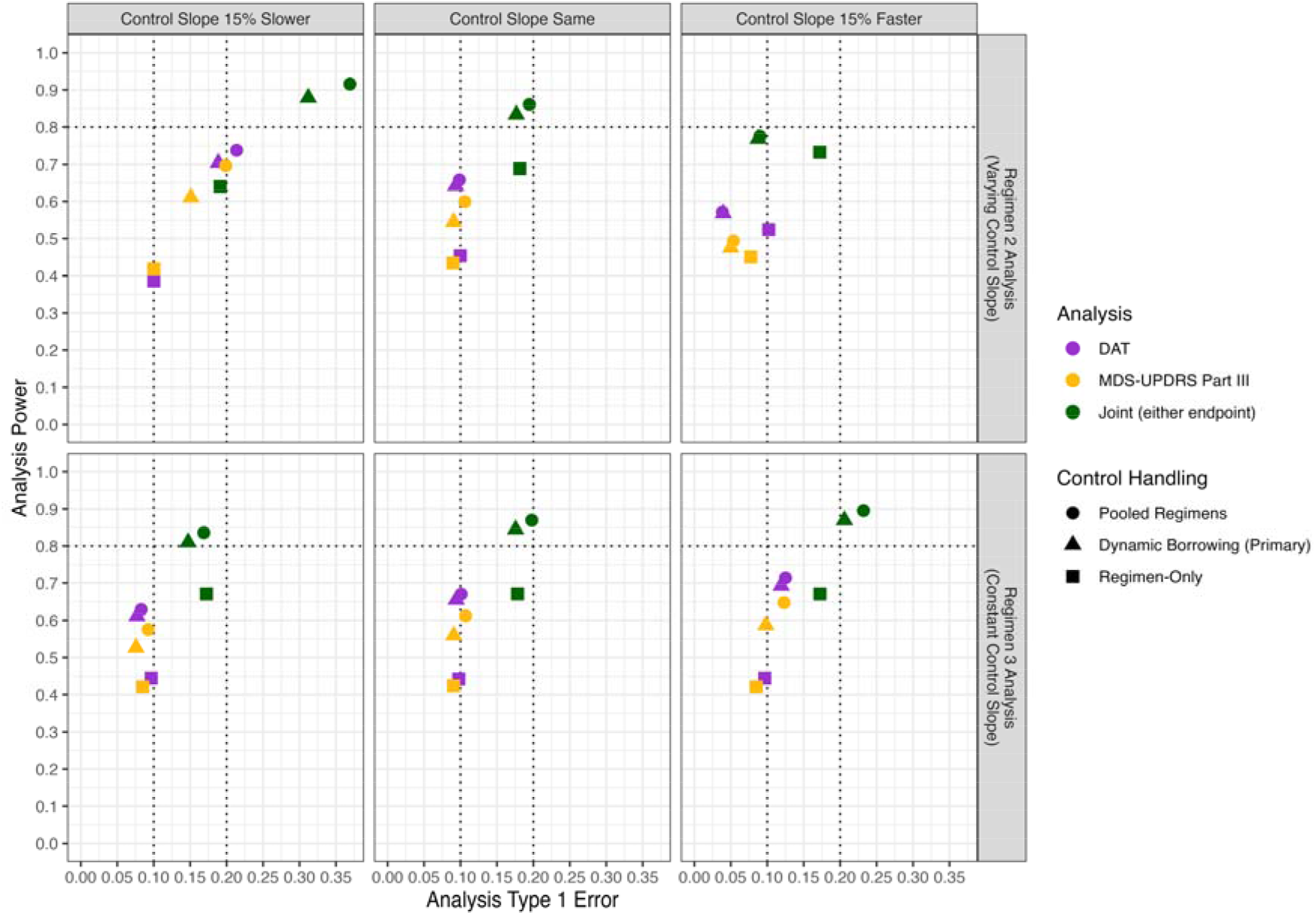
Handling control borrowing. Each panel contains a scatterplot demonstrating power as a function of Type I error via simulation. Simulations to assess power and Type I error assume 40% and 0% slowing of disease progression, respectively, on both endpoints for a regimen relative to its own control rate. In each column, the average annual progression rate for regimen 2 control participants is varied compared to all other control rates in the platform. Operating characteristics are provided for regimen 2 in the top row and regimen 3 in the bottom row. Each point corresponds to one of three endpoint analyses encoded by color: DAT bilateral putamen SBR alone, MDS-UPDRS Part III alone, and either/both endpoint (joint); and to one of three strategies for handling the collective control group encoded by shape: pooling all controls with full weight, discounting extra-regimen controls through dynamic borrowing (the primary analysis in the Master Protocol), or excluding extra-analysis regimen controls (sensitivity analysis in the Master Protocol). All analyses include 100 non-randomized controls. Dotted horizontal lines are provided at 0.10 and 0.20 to indicate target values for the individual endpoint and joint 1-sided type I error, respectively. The primary analysis model for each endpoint applies different hierarchical borrowing priors by design. The MDS-UPDRS Part III model downweighs extra-regimen control information more quickly than the DAT bilateral putamen SBR model, when the extra-regimen data progression rates disagree.

Two extreme analysis methods are presented in the figure to bracket the trade-off. Full pooling of all control data maximizes power but can inflate the individual endpoint Type I error rate in the presence of heterogeneous control progression. Restricting each analysis to regimen-specific control data maintains one-sided Type I error rates near the nominal level of 10% but reduces power. Dynamic borrowing sits between the two extremes. If the regimen 2 control progression rate is 15% slower, on average, than the rest of the platform control data sources (left column of figure), full pooling can increase the Type I error rate for regimen 2 to 0.20-0.21 on each individual endpoint because the extra-regimen control participants are progressing faster than the regimen 2-specific control participants. Dynamic borrowing discounts the discrepant extra-regimen control sources and limits the Type I error to 0.19 for the DAT bilateral putamen SBR analysis and 0.15 for the MDS-UPDRS Part III analysis. The effect on the regimen 3 analysis is conservative because inclusion of slower progressing regimen 2 control data attenuates the treatment effect estimate. The pattern is reversed when regimen 2 control participants progress more quickly (right column). The impact of differential regimen 2 control rates on regimen 3 is modest across all scenarios.

The primary endpoint analyses respond differently to the discrepancies in control rates by design. The MDS-UPDRS Part III model applies a hierarchical borrowing prior that downweighs disagreeing extra-regimen control data more rapidly than the DAT bilateral putamen SBR model because regimen-specific differences are expected to more strongly register in the clinical endpoint than the biomarker endpoint. Therefore, a stronger borrowing approach was taken for the DAT bilateral putamen SBR model. The calibration is apparent in the figure. For example, in the bottom right panel, the dynamic borrowing approach produces operating characteristics for the DAT bilateral putamen SBR endpoint that are very similar to the pooled approach (for pooled vs borrowing: 0.125 vs 0.119 Type I error, 0.714 vs 0.694 power). In contrast, the borrowing approach for MDS-UPDRS Part III is weaker, and less close to pooling (for pooled vs borrowing: 0.123 vs 0.098 Type I error, 0.648 vs 0.587 power).

### Operating characteristics

A key platform trial efficiency derives from the comparison of active treatments to the collective control group. **Figure 5** presents power for the second regimen in the platform trial. Each cell in the figure shows a range. The lower value in the range provides power for analyses that exclude non-randomized control participants (a key secondary analysis in the master protocol) and the upper value provides power including non-randomized control participants (the primary analysis). These power ranges demonstrate how dynamic borrowing of NRC participants improves joint power by up to 4%.

**Figure 5.**
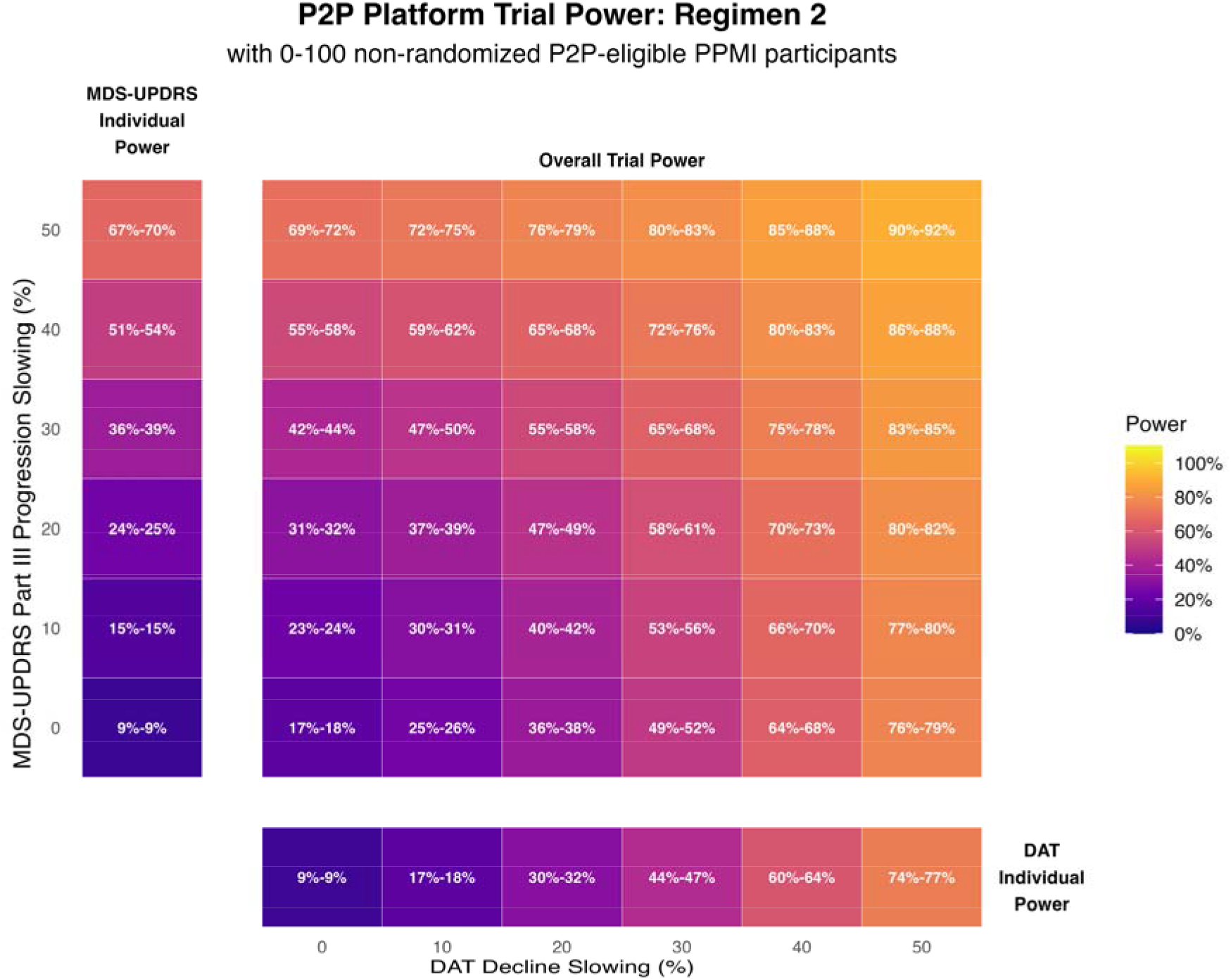
Power for P2P Platform Trial Regimen 2 Analysis. This heat map provides the trial operating characteristics for the Regimen 2 primary analysis across a range of potential treatment effects (0-50% slowing) in the DAT bilateral putamen SBR endpoint or the MDS-UPDRS Part III endpoint, as well as combinations of slowing across both endpoints. Each cell of the heat map gives the range for the overall trial power for the specified DAT bilateral putamen slowing and MDS-UPDRS Part III slowing. The lower value of this range corresponds to the power that is achieved without including the non-randomized control data in the primary analysis, while the upper value of the range corresponds to the power that is achieved when non-randomized control data from 100 participants is included in the primary analysis. The P2P platform trial primary analysis is well-powered (83%) to detect a 40% slowing on the DAT bilateral putamen endpoint and a 40% slowing on the MDS-UPDRS Part III endpoint. Power gains up to 4% occur when PPMI non-randomized control data are included in the primary analysis.

Type I error is controlled at 10% for each endpoint. A regimen can demonstrate a statistically significant treatment benefit on either endpoint to be considered successful, yielding an overall target Type I error rate of 20%. Assuming a treatment slows progression by 40% on each endpoint, we estimate 83% power for the joint primary analyses. The probability of detecting a true treatment effect of 40% slowing on DAT alone is 64%. The probability of detecting a true treatment effect of 40% slowing on MDS-UPDRS Part III alone is 54%. In individual trial simulations, observed treatment effects around 35% slowing or stronger resulted in statistically significant results for each endpoint.

Power can vary with timing of the regimen in the platform. Regimen 2 has been presented here as an example. Regimen 1 and Regimen 3 operating characteristics can be found in the Supplemental material (**Supplemental Figure S1** and **Supplemental Figure S2**). Regimen 4 is simulated to illustrate P2P’s perpetual nature and to provide realistic randomization ratios.

Because power is heavily influenced by the assumed control progression rate, we explored sensitivities to these assumptions. If the control progression rate for both DAT bilateral putamen SBR and MDS-UPDRS Part III is assumed to be 20% slower, power to detect a 40% disease-slowing treatment effect on either endpoint is 72%. On the other hand, if the control progression rates for both DAT bilateral putamen SBR and MDS-UPDRS Part III are 20% faster than expected, power to detect a 40% slowing effect is 90%. The Supplementary material provides a broader range of explored scenarios. This influence further underscores the value of the PPMI Clinical cohort registry data for enabling empirically derived simulation assumptions. Our ability to accurately predict the control progression rates on the DAT and UPDRS-III allows us to more precisely size the study for optimal Phase 2a power and efficiency.

## Discussion

P2P is the first platform clinical trial designed to test disease modifying therapeutic candidates in a biologically defined NSD Stage 2B population that precedes clinical diagnosis. This biologically defined NSD population reduces the biological heterogeneity of the population to more precisely target disease and more effectively power a clinical study. Lack of prior clinical trial data on such a population provides a challenge for a priori considerations of endpoint selection and corresponding power calculations. However, the available data and established infrastructure of the longstanding PPMI observational study addresses some of these challenges.

It is difficult to model progression in an early disease population, because there are few measurable changes that capture the disease process at that stage. PPMI data provides the largest and most robust dataset currently available to characterize the NSD population. We investigated clinical and biomarker data in biologically defined Stage 2B NSD to select endpoints with early sensitivity for phenoconversion. The data-informed recommendations for the P2P primary endpoints are based on substantial efforts to understand early and detectable changes in validated outcome assessments. Ongoing data collection and modeling efforts will continue to provide an iterative learning approach and will inform the final prespecified analysis. The MDS-UPDRS Part III endpoint was selected for statistical and clinical relevance as an early measure of manifesting disease. Selection of this clinical endpoint focuses on the motor domain but neglects non-motor diagnosis domains. Statistically, the rationale for this endpoint was based on higher sensitivity to phenoconversion and a higher signal to noise ratio. Multiple measures of non-motor dysfunction and disability are included as exploratory measures in the P2P study, and data analysis will inform which of them have appropriate performance characteristics for future studies.

The design of the P2P platform trial provides several statistical efficiencies that benefit trial participants while maintaining scientific rigor. Within the platform setting, the shared placebo control group and K:1 randomization scheme reduces the number of participants assigned toplacebo control while maintaining a 1:1 allocation ratio of active treatment to shared concurrent placebo controls to ensure well-powered analyses. The longitudinal primary analysis method leverages all available participant data, which increases power relative to other common methods such as a change from baseline analysis. The Bayesian primary analysis method facilitates dynamic borrowing of the different control groups comprising the collective control (including non-randomized control data from the PPMI study), which increases the effective sample size of the study. Bayesian modeling provides an intuitive, flexible framework that both handles regimen differences and elegantly borrows from the non-randomized cohort.

PPMI provided simulation and design-enabling longitudinal data that greatly improves the accuracy and precision of assumptions made when powering and sizing regimens for evaluation in P2P. The trial design process was highly iterative and allowed the P2P design team to fine tune design features, including the dynamic borrowing framework that underpins the handling of regimen and non-randomized cohort data in the primary analysis. Additional decisions related to understanding accrual assumptions, accumulation of trial data over time, and timing of regimens entering and exiting the platform. Clinical trial success often hinges not only on therapeutic efficacy but also on realistic assumptions about participant outcomes to ensure the detection of an effective treatment.

As an embedded trial, P2P gains statistical efficiencies from non-randomized control data. Statistical methods are well-established and have been used to borrow from historical and external control data, including in platform trials.^34^ Beyond methodological arguments, regulatory guidance has been issued on considerations for the use of external controls in clinical trials.^35^ With the infrastructure provided by PPMI, we expected the borrowed control data in P2P to have substantial similarity to the randomized control data, providing a compelling use case for borrowing.^36^ In difficult to enroll populations, this is an innovative and important supplement to randomized trial data. The PPMI observational study also provides a robust operational framework. Embedding P2P within PPMI provides operational efficiencies related to accrual and data collection, in addition to the discussed statistical efficiencies.

Despite the wealth of natural history data used to inform the P2P design, some limitations do exist. The trial population could progress at different than expected rates, and assumptions on accrual and dropout may deviate from those considered during trial design. For example, accrual is expected to be quick for participants already enrolled in the PPMI observational study at the start of P2P, followed by a slower enrollment rate after the initial bolus. Trial operating characteristics were calculated via simulation across a range of possibilities to mitigate these risks.

P2P is a learning platform that aims to assess potential disease-modifying therapies and increase knowledge across several trial dimensions. P2P is a testing ground for embedded trials and the advantages that we may gain from non-randomized cohort data. We expect to gain rich outcome data on biomarkers, clinical outcomes, including digital-health technologies. Additionally, we will learn about similarities and differences between natural history cohort accrual and retention compared to that in a randomized setting. Importantly, P2P provides a unique opportunity for early-stage intervention in the pathological cascade associated with Parkinson’s disease and other a-synucleinopathies. With robust evidence-based assumptions underpinning its design and innovative embedding within the established PPMI infrastructure, P2P is uniquely poised to test therapeutic candidates in early NSD and accelerate therapeutic development in the Parkinson’s disease space.

## Supporting information

PPMI Study Group Authors

## Data availability

Data used in the preparation of this article were obtained on 2025-09-29 from the Parkinson’s Precision Medicine Initiative (PPMI) database (https://www.ppmi-info.org/access-data-specimens/download-data), RRID:SCR_006431. For up-to-date information on the study, visit http://www.ppmi-info.org.

Protocol information for The Parkinson’s Precision Medicine Initiative (PPMI) Clinical - Establishing a Deeply Phenotyped PD Cohort can be found on protocols.io or by following this link: https://dx.doi.org/10.17504/protocols.io.n92ldmw6ol5b/v2.

This analysis was conducted by the PPMI Statistics Core and used actual dates of activity for participants, a restricted data element not available to public users of PPMI data (Tier 4 Data).

Statistical analysis codes used to perform the analyses in this article are shared on Zenodo 10.5281/zenodo.23512956.

## Acknowledgements

PPMI – a public-private partnership – is funded by the Michael J. Fox Foundation for Parkinson’s Research and funding partners, including AbbVie, Alamar Biosciences, Aligning Science Across Parkinson’s (ASAP), Arrowhead Pharma, Arvinas, AskBio, BIAL, BioArctic, Biohaven, BlueRock Therapeutics, Bristol Myers Squibb, Calico Labs, Capsida Biotherapeutics, Critical Path Institute, DaCapo Brainscience, Denali, Edmond J. Safra Foundation, Eli Lilly, Gain Therapeutics, GE Healthcare, Genentech, GSK, Insitro, Johnson & Johnson Innovative Medicine, Lundbeck, Merck, Neumora, Neuron23, Novartis, Olink, Regeneron, Roche, Sanofi, Tenvie, UCB, Vanqua Bio, Voyager Therapeutics, The Weston Family Foundation.

## Supplementary material

Supplementary material is available at *Brain* online.

