## Supplement Materials for "Statistical design of Path to Prevention platform trial in early neuronal alpha-synuclein disease"

### PPMI Observational Longitudinal Study Analysis

We analyzed data from the PPMI Clinical cohort to characterize expected endpoint behavior and generate realistic clinical trial simulations.

To characterize untreated NSD progression, we analyzed 275 PPMI Prodromal participants from the PPMI Clinical cohort who met P2P eligibility criteria. Among these P2P-eligible participants, we recorded 671 DAT bilateral putamen and 731 MDS-UPDRS Part III observations, spanning 0.7 to 3.4 years of follow-up, with a mean duration of 1.8 years.

To generate realistic endpoint data, assumptions for the untreated progression in DAT bilateral putamen SBR and MDS-UPDRS Part III are based on endpoint-specific age- and sex-adjusted linear mixed-effects models with random slope and intercept fit to longitudinal data from the 275 P2P-eligible participants. On average, DAT bilateral putamen SBR measures declined and MDS-UPDRS Part III measures increased over time, with baseline defined as the first time a participant was eligible for P2P. We estimated the average annual change using the described linear mixed-effects models. To estimate mean values at the first time of eligibility and data variability, we computed cohort means and standard deviations. Observed variability remained constant across time for DAT bilateral putamen SBR but increased as a function of time for MDS-UPDRS Part III. This difference is expected, as MDS-UDPRS Part III values move away from the measure’s floor of zero, while DAT scan measures do not approach their minimum value. Data-derived estimates on mean behavior and associated variability were used to generate accurate clinical trial simulations (**Supplemental** **Table S1)**.

To accurately characterize and simulate the multiple primary endpoints, we explored within-endpoint and across-endpoint correlations over time. As expected, endpoint values were positively correlated within each participant, with proximal times mostly showing higher correlations than more distal ones. For DAT bilateral putamen SBR, these correlations ranged from 0.8 for baseline and year one to 0.51 for baseline and year three. Within-participant MDS-UPDRS Part III correlations ranged more widely across time points (0.19 to 0.80). The DAT bilateral putamen biomarker and MDS-UPDRS Part III clinical endpoint were negatively correlated across time (**Supplemental Table S2**).

Because power is dependent on the assumed control progression rate, we explored sensitivities to our control progression rate assumptions. If both endpoints’ control progressions are slowed by 20%, we simulate the primary analysis power of 72% to detect a 40% disease-slowing treatment effect (an 11-percentage-point decrease from the 83% power in our base case). On the other hand, if both endpoints’ control progressions are 20% faster, we simulate the same primary analysis power at 90% (an increase of 7 percentage points from our base case). **Supplementary Table S3** provides a broader range of explored sensitivity scenarios.

Power varies as a function of regimen timing in the platform. Results for Regimen 2 operating characteristics are provided in Figure 4 of the main manuscript. Results for Regimen 1 and Regimen 3 operating characteristics can be found in **Supplementary Figure S1** and **Supplementary Figure S2**. These simulations assume regimens enter 0, 0.5, 1, and 1.8 years after the platform launches. Overall trial power uses a one-sided alpha of 0.20. Power for each individual endpoint uses a one-sided alpha of 0.10. Regimen 1 power for the primary analysis is 75%, which is slightly lower than Regimen 2’s power of 83% in the base case. This is primarily because of less accumulated shared control data. Regimen 3 power for the primary analysis is 84%, which is slightly higher than the power for Regimen 2, and this largely results for more accumulated data at the time of the primary analysis.

**Supplementary Tables**

**Table S1 Observed baseline value and annual change for primary endpoints**

|  | **PPMI Eligible Cohort Estimates** | | |
| --- | --- | --- | --- |
| **Primary Endpoint** | **Observed Mean at First Eligibility** | **Annual Change** | **Data-Level Standard Deviation** |
| DAT bilateral putamen SBR | 0.95 | -0.03 | 0.21 |
| MDS-UPDRS Part III | 3.44 | 1.33 | 3.94, 5.41, 6.27, 8.22^b^ |

^a^ based on September 2025 data cut

^b^ While DAT variability remained constant across years of follow-up, MDS-UPDRS III variability increased across follow-up. For this endpoint, standard deviations are provided for years 0, 1, 2, and 3.

**Table S2 Assumed Correlation between Primary Endpoints**

|  | UPDRS 0 | UPDRS 1 | UPDRS 2 | UPDRS 3 | DAT 0 | DAT 1 | DAT 2 | DAT 3 |
| --- | --- | --- | --- | --- | --- | --- | --- | --- |
| UPDRS 0 | 1 | 0.63 | 0.44 | 0.19 | -0.25 | -0.25 | -0.25 | -0.25 |
| UPDRS 1 | 0.63 | 1 | 0.68 | 0.74 | -0.25 | -0.25 | -0.25 | -0.25 |
| UPDRS 2 | 0.44 | 0.68 | 1 | 0.80 | -0.25 | -0.25 | -0.25 | -0.25 |
| UPDRS 3 | 0.19 | 0.74 | 0.80 | 1 | -0.25 | -0.25 | -0.25 | -0.25 |
| DAT 0 | -0.25 | -0.25 | -0.25 | -0.25 | 1 | 0.80 | 0.64 | 0.51 |
| DAT 1 | -0.25 | -0.25 | -0.25 | -0.25 | 0.80 | 1 | 0.80 | 0.64 |
| DAT 2 | -0.25 | -0.25 | -0.25 | -0.25 | 0.64 | 0.80 | 1 | 0.8 |
| DAT 3 | -0.25 | -0.25 | -0.25 | -0.25 | 0.51 | 0.64 | 0.8 | 1 |

We use observed correlations in past P2P-eligible PPMI natural history participants to define correlation within endpoints across time and between primary endpoints.

^a^ based on September 2025 data cut

**Table S3: Sensitivity to Slopes Table**

| DAT treatment effect  (% slowing) | MDS-UPDRS-III treatment effect (% slowing) | Adjustment to DAT Control Slope | Adjustment to MDS- UPDRS Part III Control Slope | Regimen 2 Power |
| --- | --- | --- | --- | --- |
| 30 | 30 | 0.8 | 0.8 | 0.587 |
| 30 | 30 | 1.2 | 1.2 | 0.75 |
| 30 | 40 | 0.8 | 0.8 | 0.631 |
| 30 | 40 | 1.2 | 1.2 | 0.835 |
| 40 | 30 | 0.8 | 0.8 | 0.623 |
| 40 | 30 | 1.2 | 1.2 | 0.842 |
| 40 | 40 | 0.8 | 0.8 | 0.718 |
| 40 | 40 | 1.2 | 1.2 | 0.896 |

Primary analysis power simulations under variation in the expected controls progression rates for DAT and MDS-UPDRS III measures.

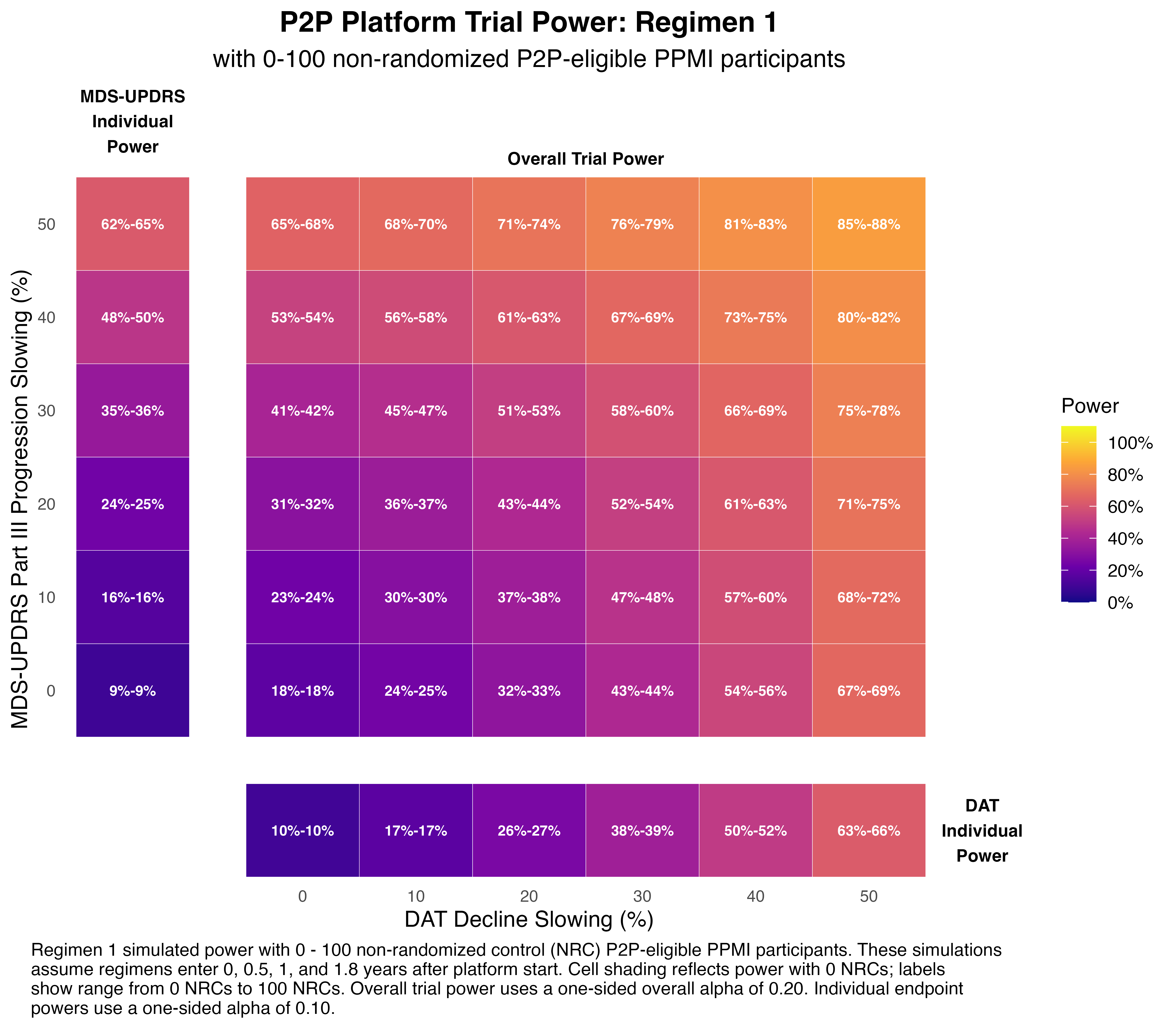
**Supplemental Figure S1 Power for P2P Platform Trial Regimen 1 Analysis.** This heat map provides the trial operating characteristics for the Regimen 1 primary analysis across a range of potential treatment effects (0-50% slowing) in the DAT bilateral putamen SBR endpoint or the MDS-UPDRS Part III endpoint, as well as combinations of slowing across both endpoints. Each cell of the heat map gives the range for the overall trial power for the specified DAT bilateral putamen SBR slowing and MDS-UPDRS Part III slowing. The lower value of this range corresponds to the power that is achieved without including the non-randomized control data in the primary analysis, while the upper value of the range corresponds to the power that is achieved when non-randomized control data from 100 participants is included in the primary analysis. The P2P platform trial primary analysis is 75% powered to detect a 40% slowing on the DAT bilateral putamen SBR endpoint and a 40% slowing on the MDS-UPDRS Part III endpoint. This is compared to the 83% power for the Regimen 2 primary analysis. The difference in power is primarily due to smaller accumulation of control data at the time of the Regimen 1 analysis.

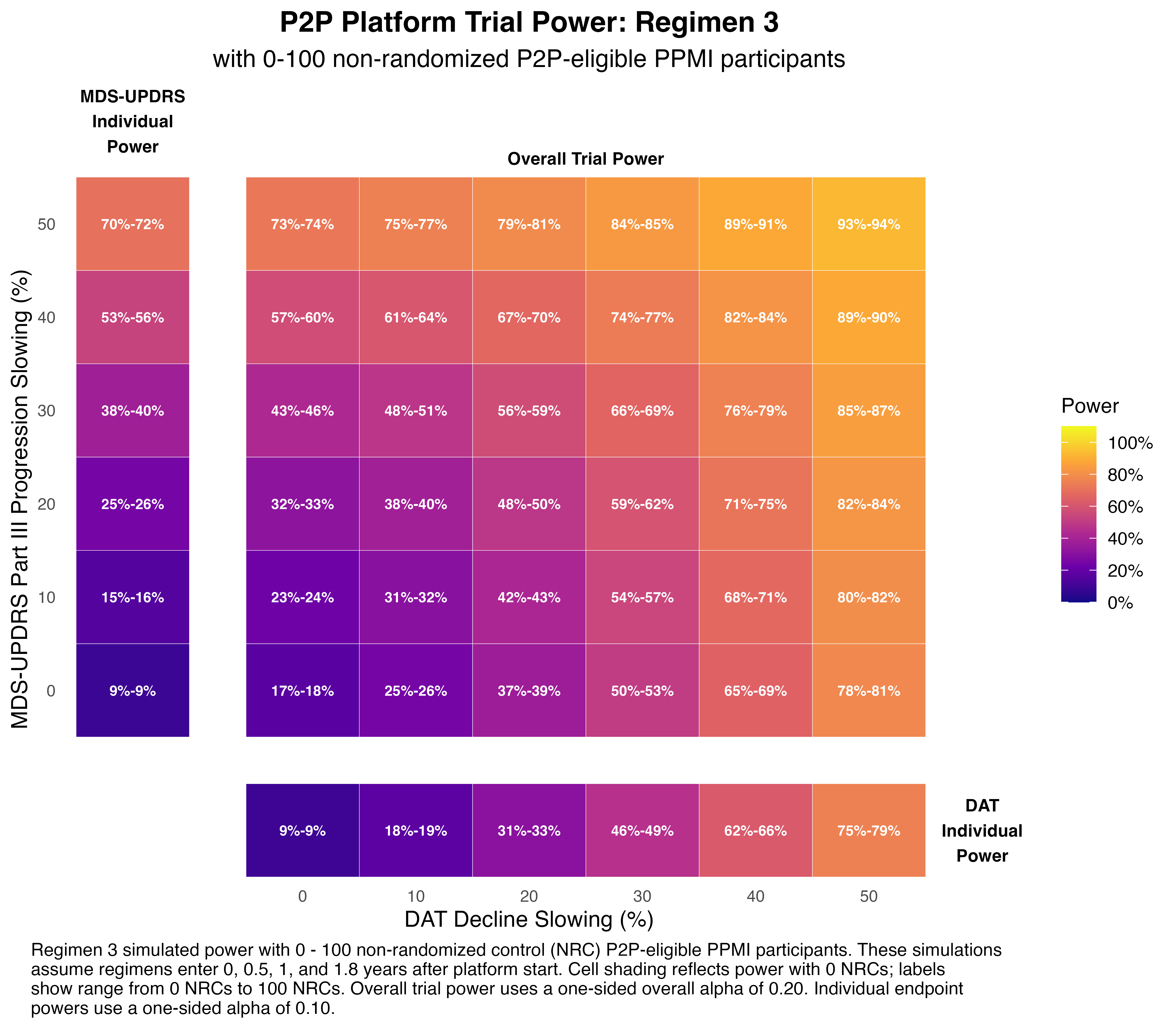
**Supplemental Figure S2 Power for P2P Platform Trial Regimen 3 Analysis.** This heat map provides the trial operating characteristics for the Regimen 1 primary analysis across a range of potential treatment effects (0-50% slowing) in the DAT bilateral putamen SBR endpoint or the MDS-UPDRS Part III endpoint, as well as combinations of slowing across both endpoints. Each cell of the heat map gives the range for the overall trial power for the specified DAT bilateral putamen SBR slowing and MDS-UPDRS Part III slowing. The lower value of this range corresponds to the power that is achieved without including the non-randomized control data in the primary analysis, while the upper value of the range corresponds to the power that is achieved when non-randomized control data from 100 participants is included in the primary analysis. The P2P platform trial primary analysis is 84% powered to detect a 40% slowing on the DAT bilateral putamen SBR endpoint and a 40% slowing on the MDS-UPDRS Part III endpoint. This is compared to the 83% power for the Regimen 2 primary analysis. The difference in power is primarily due to a larger accumulation of control data at the time of the Regimen 3 analysis.
