## Supplementary material for "Statistical design of Path to Prevention platform trial in early neuronal alpha-synuclein disease": PPMI Study Group Authors

Parkinson's Precision Medicine Initiative

### PPMI STUDY COMMITTEES, CORES, AND COLLABORATORS

#### PPMI EXECUTIVE STEERING COMMITTEE

Kenneth Marek, MD[1] (Principal Investigator); Tanya Simuni, MD[2]; Andrew Siderowf, MD[3]; Caroline M. Tanner, MD[4]; Thomas F. Tropea, DO[1]; Tatiana Foroud, PhD[5]; Lana M. Chahine, MD[4]; Brit Mollenhauer, MD[6]; Kalpana Merchant, MD[2]; Douglas Galasko, MD[7]; Christopher S. Coffey, PhD[8]; Kathleen L. Poston, MD[9]; Roseanne D. Dobkin, PhD[10]; Ethan G. Brown, MD[4]; Roy N. Alcalay, MD[11]; Dan Weintraub, MD[3]; Emily Flagg, BA[1]; Kimberly Fabrizio, BA[1]; Craig Stanley, PhD[1]; Bridget A. McMahon, BS[1]

#### PPMI STEERING COMMITTEE

Susan Bressman, MD[12]; Cornelis Blauwendraat, PhD[13]; Sonya Dumanis, PhD[13]; Paola Casalin, PhD[14]; Raymond James, RN[15]; Karl Kieburtz, MD[16]; Sneha Mantri, MS[17]; Werner Poewe, MD[18]; Michael Schwarzschild, MD[19]; John Seibyl[1], MD; David Standaert, PhD[20]; Duygu Tosun-Turgut, PhD[4]

#### MICHAEL J. FOX FOUNDATION

Sohini Chowdhury, MA[21]; Jamie Eberling, PhD[21]; Mark Frasier, PhD[21]; Leslie Kirsch, EdD[21]; Katie Kopil, PhD[21]; Maggie Kuhl, BA[21]; Alyssa O'Grady, BA[21]; Todd Sherer, PhD[21]; Tawny Willson, MBS[21]

#### PPMI STUDY CORES

*Project Management Core:* Emily Flagg, BA[1]
*Site Management Core:* Thomas F Tropea, DO[1]; Bridget McMahon, BS[1]
*Data Strategy and Technical Operations:* Craig Stanley, PhD[1]; Kim Fabrizio, BA[1]
*Data Management Core:* Dixie Ecklund, MBA[8], MSN
*Screening Core:* Tatiana Foroud, PhD[5]; Laura Heathers, BA[5]; Christopher Hobbick, BSCE[5]; Gena Antonopoulos, BSN[5]
*Imaging Core:* John Seibyl, MD[1]; Kathleen Poston, MD[9]
*Statistics Core:* Christopher Coffey, PhD[9]; Chelsea Caspell-Garcia, MS[9]; Michael Brumm, MS[9]
*Bioinformatics Core:* Arthur Toga, PhD[22]; Karen Crawford, MLIS[22]
*Biorepository Core:* Tatiana Foroud, PhD[5]; Abigail Erickson, BS[5]
*Biologics Review Committee:* Brit Mollenhauer, MD[6]; Doug Galasko, MD[7]; Kalpana Merchant, MD[2]
*Genetics Core:* Andrew Singleton, PhD[24]
*Pathology Core:* Tatiana Foroud, PhD[5]; Dirk Keene, MD[5]
*Found:* Caroline Tanner, MD[4]; Ethan Brown, MD[4]
*PPMI Online:* Carlie Tanner, MD[4]; Ethan Brown, MD[4]; Lana Chahine, MD[4]; Roseann Dobkin, PhD[10]; Monica Korell, MPH[4]

#### PPMI SITE INVESTIGATORS

Neha Prakash MD[1]; Tanya Simuni, MD[2]; Nabila Dahodwala MD[3]; Caroline Tanner, MD[4]; Brit Mollenhauer MD[7]; Susana Schnell MD[6]; Douglas Galasko, MD[7]; Anat Mirelman PhD[11]; Roy Alcalay MD[11]; Katherine Leaver MD[12]; Marie Saint-Hilaire MD[15]; Ruth Schneider MD[16]; Christopher Tarolli MD[16]; Werner Poewe, MD[18]; Corinne Horlings, MD[18]; Aleksandar Videnovic MD[19]; David Standaert PhD[20]; Marissa Dean, MD[20]; Abigail Olsen MD[24]; Rejko Krueger MD[25]; Stewart Factor DO[26]; Joseph Quinn MD[26]; Robert Hauser MD[28]; Amy Amara PhD[29]; Michelle Fullard MD[29]; Cyrus Zabetian MD[30]; Hubert Fernandez MD[31]; Kathrin Brockmann MD[32]; Isabel Wurster PhD[32]; Yen Tai PhD[33]; Paolo Barone PhD[34]; Marina Picillo MD[34]; Stuart Isaacson MD[35]; Alberto Espay MD[36]; Eduardo Tolosa PhD[37]; Alicia Garrido MD[37]; Javier Ruiz Martinez PhD[38]; Leonidas Stefanis PhD[39]; Christos Koros MD[39]; Kelvin Chou MD[40]; Lorraine Kalia MD[41]; Connie Marras PhD[41]; David Grimes MD[42]; Tiago Mestre PhD[42]; Rajesh Pahwa MD[43]; Mark Lew MD[44]; Holly Shill MD[45]; Shyamal Mehta MD[46]; Giulietta Riboldi MD[47]; Nikolaus McFarland PhD[48]; Ron Postuma MD[49]; Zoltan Mari MD[50]; David Ledingham MD[51]; Nicola Pavese PhD[51]; Michele Hu PhD[52]; Norbert Brüggemann MD[53]; Christine Klein MD[53]; Bastiaan Bloem PhD[54]; Cristina Simonet PhD[55]; Alastair Noyce PhD[55]; Anette Janzen PhD[56]; Wolfgang Oertel PhD[56]; Njideka Okubadejo MD[57]; David Shprecher DO[58]; Arjun Tarakad MD[59]; Emile Moukheiber MD[60]

#### PPMI SITE COORDINATORS

Joy Anatala[1]; Robert Moss[1]; Mackenzee George[1]; Kristen Hagwell[1]; Ashley Romano[1]; Sarah Chatfield[1]; Stephanie Roman[1]; Carla Aranda[2]; Karen Williams[2]; Sophia Melton[2]; Karina Benson[2]; Sarah Rogers[2]; Ritikha Vengadesh[3]; Joana Salievska[3]; Adaeze Njoku[3]; Taylor Siegal[3]; Jaime Heller[4]; Primi Ranola[4]; Farah Kausar[4]; Jacque Perkins[4]; Diana Willeke[6]; Elizabeth Kalinkara-Gomez[6]; Susanne Scaglione[6]; Janelle Rodriguez[7]; Nobuko Kemmotsu[7]; May Eshel[11]; Deborah Raymond[12]; Emma Margolis[12]; Ivy Gu[12]; Maya Rawal[12]; Abigail Desrosiers[15]; Raymond James[15]; Lauren Jackson[16]; Brittany Clark[16]; Iris Egner[18]; Nathan Fogarty[19]; Courtney Blair[20]; Lauren Ruffrage[20]; Sherri Mosovsky[24]; Berenice Sevilla[25]; Sonja Jonsdottir[25]; Claire Pauly[25]; Emna Bouhajja[25]; Barbara Sommerfeld[26]; Madison Christopher[27]; Montana Pearson[28]; Erica Botting[28]; Gabriella Mazur[28]; Danielle Derlein[29]; Evan Doll[29]; Ying Liu[29]; Nathan Cheng[29]; Laura Gorla[29]; Lucas Lattanzio[29], Sabrina Akhter Mim[29]; Dillan Patel[29]; Ciera Cobb[30]; Gianna Dal Porto[30]; Jennifer Mule[31]; Stuart Houltham[31]; Renee Dancie[31]; Ella Hilt[32]; Aldazier Jakiran[33]; Syeda Visha Fatima[33]; Dominga Valentino[34]; Lisbeth Pennente[35]; Bobbie Stubbeman[36]; Ayesha Shaik[36]; Katherin Galvez[36]; Valeria Ravasi[37]; Maria Fernanda Farfan Ale[37]; Ioana Croitoru[38]; Nikolas Papagiannakis[39]; Evangelia Stanitsa[39]; Frank Ferrari[40]; Mengyu Zheng[41]; Shawna Reddie[42]; Alicia Alejandra[43]; Sydney Chappell[43]; Carsyn Smith[43]; Colette Worthington[43]; Alejandra Valenzuela[44]; Aliana Valenzuela[44]; Bethzaida Herrera[45]; Martin Salamanca Jr.[45]; Sara Dresler[46]; Neil Santos[46]; Fahrial Esha[47]; Danayeat Abraha[47]; Mark Belio[47]; Kyle Rizer[48]; Nadine Zablith[49]; Roxana Garcia[50]; Liliana Dumitrescu[50]; Victoria Kate Foster[51]; Debra Galley[51]; Robyn Iredale[51]; Jamil Razzaque[52]; Madita Grümmer[53]; Lian Feenstra[54]; Natalie Donkor[55]; Ayat Azhar[55]; Rhiannon Laban[55]; Ruby Lathey[55]; Anisa Shahid[55]; Elisabeth Sittig[56]; Oluwadamilola Ojo[57]; Kelly Clark[58]; Rory Mahabir[59]; Giselle Silva[59]; Ramya Palani[60]; Shamera Willoughby[60]

**INSTITUTIONS AND AFFILIATIONS**

1. Institute for Neurodegenerative Disorders; New Haven, CT, USA
2. Northwestern University; Chicago, IL, USA
3. University of Pennsylvania; Philadelphia, PA, USA
4. University of California, San Francisco; San Francisco, CA, USA
5. Indiana University; Indianapolis, IN, USA
6. Paracelsus-Elena Klinik; Kassel, Germany
7. University of California, San Diego, San Diego, CA, USA
8. University of Iowa; Iowa City, IA, USA
9. Stanford University; Stanford, CA, USA
10. Rutgers University; New Brunswick, NJ, USA
11. Tel Aviv Sourasky Medical Center; Tel Aviv, Israel
12. Mount Sinai Beth Israel; New York, NY, USA
13. Coalition for Aligning Science; Chevy Chase, MD, USA
14. BioRep; Milan, Italy
15. Boston University School of Medicine; Boston, MA, USA
16. University of Rochester; Rochester, NY, USA
17. Duke University; Durham, NC, USA
18. University of Innsbruck; Innsbruck, Austria
19. Massachusetts General Hospital; Boston, MA, USA
20. University of Alabama at Birmingham; Birmingham, AL, USA
21. The Michael J. Fox Foundation for Parkinson's Research; New York, NY, USA
22. Laboratory of Neuroimaging (LONI), USC; Los Angeles, CA, USA
23. National Institute on Aging, NIH; Bethesda, MD, USA
24. University of Pittsburgh; Pittsburgh, PA, USA
25. University of Luxembourg; Esch-sur-Alzette, Luxembourg
26. Emory University; Atlanta, GA, USA
27. Oregon Health and Science University; Portland, OR, USA
28. University of South Florida; Tampa, FL, USA
29. University of Colorado; Aurora, CO, USA
30. VA Puget Sound Health System; Seattle, WA, USA
31. Cleveland Clinic; Cleveland, OH, USA
32. University of Tubingen; Tubingen, Germany
33. Imperial College of London; London, UK
34. University of Salerno; Salerno, Italy
35. Parkinson's Disease and Movement Disorders Center; Boca Raton, FL, USA
36. University of Cincinnati; Cincinnati, OH, USA
37. Hospital Clinic of Barcelona; Barcelona, Spain
38. Hospital Universitario Donostia; San Sebastian, Spain
39. University of Athens; Athens, Greece
40. University of Michigan; Ann Arbor, MI, USA
41. Toronto Western Hospital; Toronto, Canada
42. The Ottawa Hospital; Ottawa, Canada
43. University of Kansas Medical Center; Kansas City, KS, USA
44. Keck School of Medicine of the University of Southern California; Los Angeles, CA, USA
45. Barrow Neurological Institute; Phoenix, AZ, USA
46. Mayo Clinic Arizona; Scottsdale, AZ, USA
47. NYU Langone Medical Center; New York, NY, USA
48. University of Florida; Gainesville, FL, USA
49. Montreal Neurological Institute and Hospital/McGill; Montreal, QC, Canada
50. Cleveland Clinic-Las Vegas Lou Ruvo Center for Brain Health; Las Vegas, NV, USA
51. Clinical Ageing Research Unit; Newcastle, UK
52. John Radcliffe Hospital Oxford and Oxford University; Oxford, UK
53. University of Luebeck; Luebeck, Germany
54. Radboud University; Nijmegen, Netherlands
55. Queen Mary University of London; London, UK
56. Philipps-University Marburg; Marburg, Germany
57. University of Lagos; Lagos, Nigeria
58. Banner Sun Health Research Institute; Sun City, AZ, USA
59. Baylor College of Medicine; Houston, TX, USA
60. Johns Hopkins University; Baltimore, MD, USA

**PPMI STUDY FUNDING PARTNER STATEMENT**

PPMI – a public-private partnership – is funded by the Michael J. Fox Foundation for Parkinson's Research and funding partners, including AbbVie, Alamar Biosciences, Aligning Science Across Parkinson's (ASAP), Arrowhead Pharma, Arvinas, AskBio, BIAL, BioArctic, Biohaven, BlueRock Therapeutics, Bristol Myers Squibb, Calico Labs, Capsida Biotherapeutics, Critical Path Institute, DaCapo Brainscience, Denali, Edmond J. Safra Foundation, Eli Lilly, Gain Therapeutics, GE Healthcare, Genentech, GSK, Insitro, Johnson & Johnson Innovative Medicine, Lundbeck, Merck, Neumora, Neuron23, Novartis, Olink, Regeneron, Roche, Sanofi, Tenvie, UCB, Vanqua Bio, Voyager Therapeutics, The Weston Family Foundation.
